# Effects and dynamics of D-alanine in diabetes

**DOI:** 10.1101/2024.12.22.24319080

**Authors:** Shinsuke Sakai, Hiroki Okushima, Yukimasa Iwata, Terumasa Hayashi, Shiro Takahara, Masaru Horio, Masashi Mita, Maiko Nakane, Shihoko Kimura-Ohba, Ryoichi Imamura, Yoichi Kakuta, Masayuki Mizui, Yoshitaka Isaka, Tomonori Kimura

## Abstract

**Background:** D-alanine, a rare enantiomer of alanine, mediates renal gluconeogenesis and protects against viral infections by regulating the circadian clock’s transcriptional network. These features of D-alanine are associated with the diabetes, which disrupts circadian rhythm and worsens Covid-19 outcomes. This study examined the effects and dynamics of D-alanine in diabetic conditions.

**Methods:** Blood and urine levels of D-alanine were measured in diabetic model mice and patients with diabetic kidney disease using a two-dimensional high-performance liquid chromatography system. Gluconeogenic activity of D-alanine was assessed by glucose production assay in *ex vivo*-cultured kidney cells. Glucose tolerance test was performed in mice treated with D-alanine.

**Results:** The circadian rhythm of D-alanine, present in healthy mice, was disrupted in diabetic model mice. Patients with diabetic kidney disease showed abnormal urinary excretion of D-alanine. In diabetic mice, D-alanine stimulated gluconeogenesis in kidneys. Although D-alanine treatment temporarily raised blood glucose levels, repetitive treatment of D-alanine did not worsen glucose tolerance in the tested conditions.

**Conclusions:** An abnormal circadian rhythm of D-alanine is a hallmark of diabetes. In diabetic mice, D-alanine affects kidney function without worsening the diabetic conditions. D-Alanine may provide a therapeutic option for diabetes by correcting circadian rhythms and treating viral infections.

## Introduction

The aberrant circadian rhythm of glucose metabolism is associated with the pathogenesis of diabetes.^1^ Among the various aspects of glucose metabolism, the enzymatic activities involved in gluconeogenesis exhibit a circadian rhythm in both kidney and liver.^2,3^ Gluconeogenesis is the process by which glucose is produced from intermediate metabolites, such as pyruvate and L-amino acids. The roles of gluconeogenesis in the kidney and liver are equally important, as they help maintain blood glucose levels, particularly during fasting conditions.^2,4^ In the kidney, gluconeogenesis occurs in the proximal tubules, where gluconeogenic enzymes, such as glucose-6-phosphatase, are abundant.^2,4^ This process is activated under diabetic conditions.^5^ Additionally, patients with diabetes are at high risk for infections.^6^ For instance, in the case of Covid-19, diabetes serves as a significant risk factor.^7^ Since immune responses are regulated by circadian rhythms,^8^ impaired circadian rhythms in a broad sense may exacerbate the severity of infections in patients with diabetes.^9^

Circadian rhythm is an innate daily cycle that regulates the activities of living organisms.^10^ Circadian rhythms modulate a variety of biological processes through a transcriptional network.^11^ The core machinery of this system is comprised of circadian clock genes.^11^ A subset of these genes, such as Bmal1 and Cry2, forms a molecular negative feedback loop that generates a transcriptional circadian rhythm. Additionally, the components of circadian clock genes also regulate circadian-related biological processes through transcription. This intricate circadian transcriptional network interacts with a diverse array of biological processes. However, a significant number of circadian clock components remain unidentified.^11^

Recently, D-alanine has been identified as a molecule that bridges circadian rhythms with various biological processes. D-Alanine, one of the D-amino acids, is a rare stereoisomer of the more abundant L-alanine. The relationship between D-alanine and circadian rhythms is both multifaceted and profound. Blood levels of D-alanine exhibit a clear circadian rhythm.^12,13^ The rhythm of D-alanine is characterized by a pronounced sinusoidal variation among known metabolites, allowing for the identification of daily peaks and troughs with relative ease. This rhythm is generated through the excretion of D-alanine via urine by the kidney, suggesting that D-alanine plays a role in kidney function. Indeed, D-alanine influences the kidney and transmits signals from the circadian rhythm. D-Alanine has been shown to reduce the severity of viral infections, such as Covid-19 and the influenzae virus, indicating another link to circadian rhythms. Treatment with D-alanine may also enhance daily rhythms in conditions where circadian rhythms are disrupted. D-Alanine may exhibit contrasting effects under diabetic conditions. On one hand, D-alanine plays a regulatory role in maintaining the circadian rhythm, which is often disrupted in patients with diabetes. On the other hand, D-alanine may elevate blood glucose levels by inducing gluconeogenesis. To clarify this paradoxical situations, we investigated the dynamics and effects of D-alanine in diabetic condition using mice models.

## Methods

### Animals

C57BL/6 mice and B6.BKS(D)-Lepr^db^/J (*db*/+ mice) were purchased from SLC (Tokyo, Japan).

For the analysis of circadian oscillations in D-alanine, male mice (7-to 10-week-old) were individually caged and housed in light-tight, ventilated closets within a temperature- and humidity-controlled facility. Mice were entrained on a 12-h light:12-h dark (LD) cycle for 2 weeks, and blood samples were collected from trunk after decapitation. Urine samples were collected through bladder puncture.

For the intraperitoneal injection, mice starved for 6 h were injected with D-alanine at the dose 12.5 μmoL/g. Preliminary, mice were injected with 12.5 μmoL/g of D-alanine for three times with 12-h of interval, and confirmed the sufficient increase in blood D-alanine level. Blood samples were transcardially collected from mice under anesthesia.

In the experiments with diabetic models, five-week-old mice were fed with a normal diet (12.8% of kcal from fat: 5% fat, 23% protein, and 55% carbohydrate) or a high-fat diet (HFD 62.2% of kcal from fat: 35% fat, 23% protein, and 25% carbohydrate) (Oriental Yeast, Osaka, Japan) for 2 months. Then, diabetic mice were treated with 0.5% D-alanine via a water bottle for 2 weeks and 8 weeks.

Separately, *db*/*db* mice fed with normal diet or C57BL/6 mice fed with HFD, were starved for 6 h, followed by the intraperitoneal injection with 12.5 μmoL/g of D-alanine once.

All animal experiments were conducted in compliance with the Guidelines for Japanese Animal Protection and Management Law and performed under protocols approved by the Animal Research Committee of Osaka University

### Glucose Tolerance Test

To mice that had been fasted for 6 hours, glucose (2 g/kg body weight, Nakalai Tescque,16806-25) was intraperitoneally injected for the glucose tolerance test. Blood samples were collected from the tail vein before administering glucose and after 30, 60, and 120 minutes. Levels of glucose in whole blood were determined using a blood glucose monitoring unit (*Glutest Sensor Neo, Sanwa Kagaku Kenkyusho*, Aichi, Japan).

### Assessment of Glucose-6-phosphatase and D-amino acid oxidase activity

Glucose-6-phosphatase activity was assessed as described previously with some modifications.^14^ Briefly, the kidney cortex was homogenized in ice-cold saline, and the supernatant was collected after centrifugation for 10 minutes at 3,000 g. 10 uL of the supernatants was mixed with 150 uL of 0.1 M malate buffer containing 200 mM glucose-6-phosphate, and were incubated at 37 °C for 10 minutes. After centrifugation for 10 minutes at 4,500 rpm, 50 uL of Taussky Short Color Reagent (ammonium molybdate solution with ferrous sulfate) was applied. The amount of inorganic phosphate liberated from glucose-6-phosphate in the reaction solution was measured by spectroscopy with the absorbance at 660 nm.

### *ex vivo* glucose production assay

Proximal tubular epithelial cells (PTECs) were isolated from mice as described previously.^15^ After anesthetization and perfusion with saline, the kidneys from mice were immediately removed and placed in cold phosphate buffer solution (PBS) (4 °C). After the renal capsule was removed, the kidney was cut sagittally, and the cortex was finely minced and transferred to Hanks’ Balanced Salt Solution containing 0.25mg/ml Liberase (Roche, Penzberg, Germany) for 30 minutes at 37 °C with gentle shaking. After digestion, suspended cells were washed in glucose-free HEPES buffer consisted of 5.4 mM KCl, 143 mM NaCl, 1.8 mM CaCl_2_, 0.9mM NaH_2_PO_4_, 0.8 mM MgSO_4_, and 5 mM HEPES buffer, pH 7.4, for 30 minutes at 37 °C with gentle shaking. Then, PTECs were incubated with glucose-free HEPES buffer containing either L- or D-alanine at 1 mM or indicated levels in the legend for 1 h. Usually, the doses of ∼10 mM of D-amino acids were selected in cell culture experiments to eliminate the effect of L-amino acids, which are present abundantly.^16,17^ In this experiment, we reduced the dose of D-alanine as we performed in L-amino acid-free medium. Glucose formation was calculated from the glucose levels in the medium measured by the glucose oxidase method using the *Amplex Red/*HRP assay kit (A22189 Invitrogen, Manassas, CA).

### Sample preparation for two-dimensional high-performance liquid chromatography

Sample preparation for two-dimensional high-performance liquid chromatography (2D-HPLC) was performed as previously described.^12,18^ In brief, 20-fold volumes of methanol were added to the sample and 10 μL of the supernatant obtained from the methanol homogenate was used for NBD derivatization (0.5 μL of the plasma was used for the reaction). After drying the solution, 20 μL of 200 mM sodium borate buffer (pH 8.0) and 5 μL of fluorescence labeling reagent (40 mM 4-fluoro-7-nitro-2,1,3-benzoxadiazole (NBD-F) in anhydrous MeCN) were added, then heated at 60°C for 2 min. An aqueous 0.1 % (v/v) trifluoroacetic acid (TFA) solution (75 μL) was added, and 2μL of the reaction mixture was subjected to 2D-HPLC.

### Determination of alanine enantiomers by 2D-HPLC

The enantiomers of alanine were quantified using the 2D-HPLC platform, as previously described,^12,18^ with the shape-fitting algorithm.^19^ Briefly, the NBD derivatives of the amino acids were separated from numerous intrinsic substances using a reversed-phase column (Singularity RP column, 1.0 mm i.d. × 50 mm; provided by KAGAMI Inc., Ibaraki, Japan) with the gradient elution using aqueous mobile phases containing MeCN and formic acid. In order to separately determine the D-and L-forms of alanine, the fraction of alanine was automatically collected using a multi-loop valve, and transferred to enantioselective column (Singularity CSP-001S, 1.5 mm i.d. × 75 mm; KAGAMI Inc.). The mobile phases are the mixed solution of MeOH-MeCN containing formic acid, and the fluorescence detection of the NBD-amino acids was carried out at 530 nm with excitation at 470 nm. The fluorescence detector utilizes two photomultiplier tubes to cover high and low ranges and enables simultaneous and accurate measurement of both abundant L-alanine and trace D-alanine in human samples. D-Alanine ratio was calculated as plasma D-alanine levels divided by the sum of L- and D-alanine levels.

### Biochemical Measurements

Plasma glucose levels were measured in 60 minutes after intraperitoneal injection of D-alanine (25 *μ*mol / kg body weight). Plasma levels of glucose were measured using the Glucose CII-test (Wako). All kits were used in accordance with the manufacturers’ protocols.

### Study population of human patients with diabetes

We prospectively enrolled diabetic patients undergoing their first kidney biopsy between 2006 and 2016 at the Department of Kidney Disease and Hypertension, Osaka General Medical Center as described previously.^20,21^ The plasma and urinary levels of D-alanine were measured from the samples before kidney biopsy.

Separately, blood samples from 60 potential living kidney transplant donors and 12 healthy volunteers were analyzed.^20^ Blood samples were obtained during daytime to reduce the effect of intraday variation. The study protocol was approved by the Ethical Committees of Osaka University (#22571). This study was conducted in compliance with the ethical principles of the Declaration of Helsinki, and all participants gave written informed consent. The clinical and research activities being reported are consistent with the Principles of the Declaration of Istanbul as outlined in the ‘Declaration of Istanbul on Organ Trafficking and Transplant Tourism’.

Fractional excretion (FE) was calculated from clearance of substrate divided by creatinine clearance, as follows:

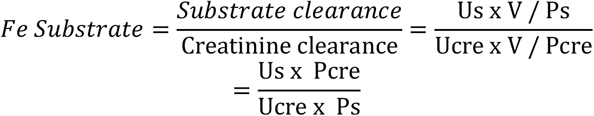

where Us and Ps represent urine and plasma levels of substrate, respectively. Fractional excretion is the ratio of a substrate filtered by the kidney glomeruli that is excreted in the urine. Serum and urine creatinine were measured enzymatically, and D-/L-serine was measured as aforementioned using the same sample. Low and high fractional excretion indicates increased tubular reabsorption and increased excretion, respectively.

### Statistics

Animals in the same litter were randomly assigned to different treatment groups. No animals or data were excluded from the analysis. Data normality was tested either by D’Agostino–Pearson omnibus normality test or by graphical test. Continuous variables are presented as means ± SEM, or as medians and ranges when indicated. Numbers of experiments are provided in the figure legends. Either two-tailed Student’s *t* test (paired or unpaired), one- or two-way ANOVA with Dunnett’s post-hoc test was used. Data were analyzed using GraphPad Prism or STATA. Statistical significance was defined as P < 0.05.

## Results

### Increased urinary excretion of D-alanine without a circadian rhythm in mouse models of diabetes

D-Alanine exhibits a distinct circadian rhythm in both blood and urine, whereas the disrupted circadian rhythm is believed to exacerbate diabetic conditions. We first investigated whether the circadian rhythm of D-alanine is affected under diabetic conditions. For this purpose, we measured the levels of D-alanine in the blood and urine of *db*/*db* mice during daytime, when levels are elevated in healthy wild type mice, and at night, when levels are reduced. In type 2 diabetes model mice characterized by obesity, plasma level of D-alanine was consistently lower than that in non-obese mice (Figure 1A). Furthermore, the circadian variation of blood D-alanine level was absent in diabetic mice.

**Figure 1.**
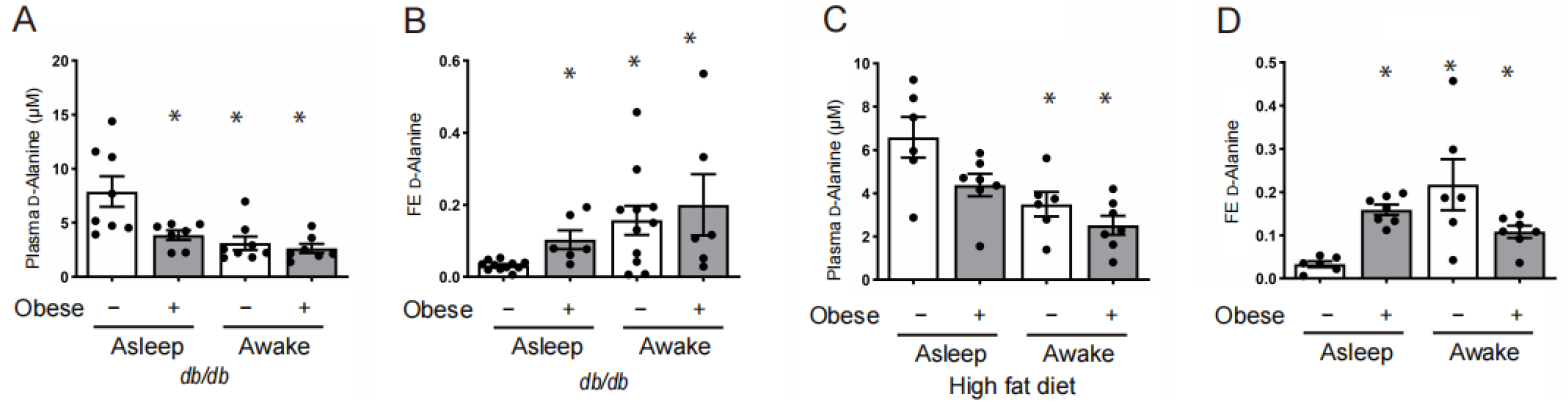
Increased urinary excretion without a circadian rhythm of D-alanine in diabetic mouse models. (A and C) Plasma level and (B and D) urinary fractional excretion (FE) of D-alanine in either non-obese or obese mice when asleep and when awake. *n* = 7−9. As obese mice, (A and B) *db*/*db* mice or (C and D) mice fed high fat diet were used. Data, means ± SEM. Statistic, one-way ANOVA \**P* < 0.05 versus healthy control during asleep.

To investigate the reason for the reduced blood level of D-alanine in diabetic mice, we examined its urinary excretion. By analysing blood and urine, we calculated the fractional excretion (FE) of D-alanine, an index of how kidney excretes D-alanine into the urine. FE of D-alanine was consistently higher in the diabetic mice compared to non-obese mice (Figure 1B). This increased urinary excretion of D-alanine likely contributes to the suppression of blood levels of D-alanine.

To verify these findings, we profiled D-alanine levels in mice fed a high-fat diet (HFD). The consistently low blood levels and elevated urinary excretion of D-alanine were confirmed in HFD-induced obese mice (Figure 1, C and D). The increased urinary excretion of D-alanine, coupled with the absence of circadian oscillation, is a indicative diabetic conditions.

### Increased urinary excretion of D-alanine in patients with type 2 diabetes

We also examined the level of D-alanine in human patients. Blood and urine samples from biopsy-proven patients with diabetic kidney disease were analyzed.^20,21^ For comparison, samples from living kidney donors were also evaluated.^22,23^ Background data indicate that diabetic patients tend to have worse kidney function (Supplementary Table 1). Patients with diabetes showed a significant increase in the FE of D-alanine (Figure 2). Although D-alanine accumulates in the blood due to decreased kidney function in these patients,^24^ the increased urinary excretion of D-amino acids despite reduced glomerular filtration is a unique phenomenon observed in patients with diabetic kidney disease.^20^

**Figure 2.**
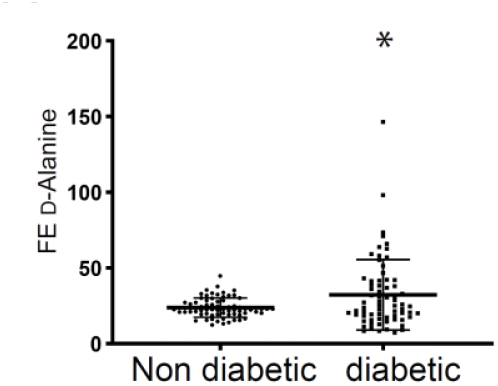
Increased urinary excretion of D-alanine in patients with type 2 diabetes. Urinary fractional excretion (FE) in human patients with diabetes. *n* = 73 for non-diabetics and 69 for diabetics. Statistics, two-tailed unpaired *t*-test. \**P* < 0.05.

### D-Alanine enhances gluconeogenesis in diabetic mouse models

Gluconeogenesis is known to be active in *db*/*db* mice and in mice fed a high-fat diet (HFD),^25,26^ while D-alanine has been shown to activate gluconeogenesis in the kidney. We investigated whether D-alanine affects gluconeogenesis under diabetic conditions. For this purpose, we employed *ex vivo* culture system to directly measure gluconeogenic activity in kidney cells, since immortalized cell lines do not possess a complete machinery necessary for gluconeogenesis.^27^ As reported, D-alanine induced glucose production in the kidney of HFD-treated mice *ex vivo* (Figure 3A).

**Figure 3.**
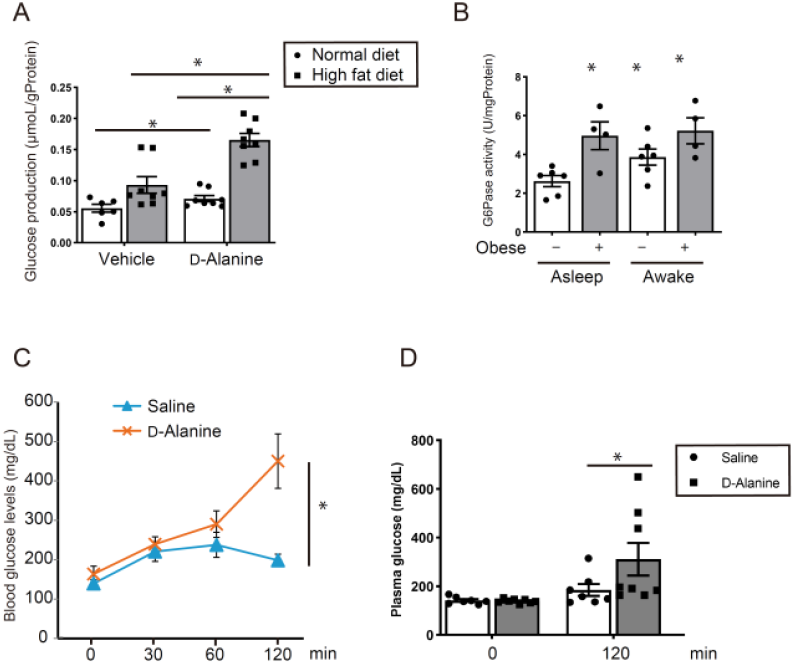
D-Alanine enhances gluconeogenesis in diabetic model mice. (A) Glucose production from D-alanine in isolated proximal tubular cells from normal diet or high fat diet-fed mice. (B) Glucose-6-phosphatase (G6Pase) activity in kidney from high fat diet (HFD)-fed mice when asleep and when awake. *n* = 7−9. (C and D) Blood glucose levels in (C) *db*/*db* mice and (D) HFD-fed mice injected with D-alanine at 0 PM. Statistic, (A, B, D) one-way ANOVA and (C) two-way ANOVA. \**P* < 0.05.

The gluconeogenic activity in diabetic mice, which is slightly elevated at baseline, was also enhanced by D-alanine. D-Alanine can augment the gluconeogenic activity in diabetic mice.

### Lost circadian oscillation of gluconeogenic activity of diabetic mice

We aimed to investigate the effects of D-alanine on gluconeogenesis under diabetic conditions. Gluconeogenic activity exhibits a circadian oscillation, and its disruption is a characteristic of diabetes. We investigated the circadian oscillation of the gluconeogenic activity in diabetic mouse model. We measured glucose-6-phosphatase (G6Pase) activity in kidney of HFD-fed mice when asleep and when awake (Figure 3B). As reported, the gluconeogenic activity in diabetic model mice was higher than that in normal mice. Importantly, this activity remained persistently elevated and lost its circadian oscillation. The loss of circadian oscillation in gluconeogenic activity is a distinctive feature of diabetic mice.

### A single dose of D-alanine treatment increases blood glucose level in diabetic mice

Since D-alanine affects gluconeogenesis even in diabetic conditions, and its administration raises blood glucose level, we investigated whether D-alanine also affects blood glucose level in diabetic conditions. For this purpose, we measured the blood glucose levels in *db*/*db* mice and in mice fed HFD that were injected intraperitoneally with D-alanine (Figure 3, C and D). The treatment with D-alanine further elevated blood glucose levels in both diabetic model groups. These findings suggest that D-alanine can activate gluconeogenesis and elevate blood glucose level even in diabetic conditions.

### Long-term treatment of D-alanine enhances glucose-related profiles in diabetic mice

D-alanine can induce gluconeogenesis even under diabetic conditions, while it may also improve diabetic states by correcting circadian rhythms. To determine the clinical significance of D-alanine under more physiological conditions, we investigated the effects of long-term D-alanine treatment in diabetic condition. Glucose tolerance test was performed on mice fed HFD following intraperitoneal injections of either saline or D-alanine for 2 weeks and 8 weeks. The results showed that repetitive treatment of D-alanine for 2 weeks did not worsen the glucose tolerance (Figure 4, A and B). Furthermore, treatment of D-alanine for 8 weeks significantly improved glucose tolerance (Figure 4, C and D). These findings suggest that long-term D-alanine treatment may enahnce glucose tolerance.

**Figure 4.**
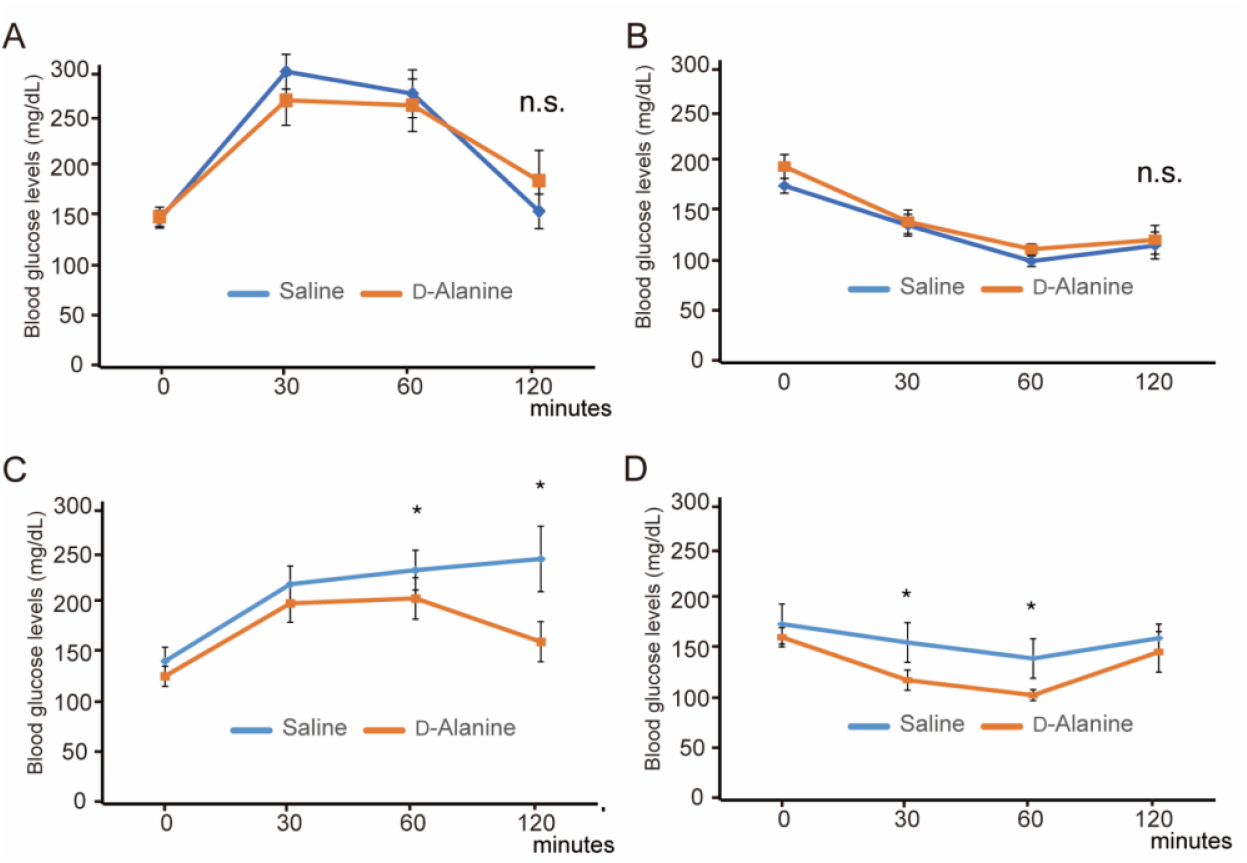
D-Alanine improves glucose profiles in diabetic mice. (A and B) (A) Glucose and (B) insulin tolerance tests in mice fed a high-fat diet (HFD) after a 2-weeks of injections of either saline or D-alanine at 8 PM. (C and D) (C) Glucose and (D) insulin tolerance tests performed after 8 weeks of the same treatment. *n* = 7−9. Statistics, two-way ANOVA. \**P* < 0.05. n.s., not significant.

## Discussion

This study elucidates the dynamics and effects of D-alanine in the context of circadian rhythm under diabetic conditions. In contrast to normal mice, the circadian rhythm of D-alanine is disrupted in diabetic mouse models, as evidently demonstrated by persistently elevated urinary excretion and reduced blood level of D-alanine. Urinary excretion profile of D-alanine is significantly elevated in patients with diabetic kidney disease. In the kidney of diabetic model mice, gluconeogenic activity is characterized by a lack of circadian rhythm and remains consistently high. Notably, even under these conditions, a single administration of D-alanine can stimulate gluconeogenesis in the kidney and elevates blood glucose level, similar to the response observed in normal mice. However, long-term treatment with D-alanine enhances glucose tolerance by restoring circadian rhythm. D-Alanine may represent a therapeutic option for diabetes by correcting circadian rhythm and reducing the severity of viral infections.

In this study, we successfully identified the disrupted circadian rhythm as a hallmark of diabetic conditions. The connection between circadian rhythm and diabetes has primarily been established through genetic studies to date. Various forms of impaired glucose metabolism, particularly glucose intolerance, have been observed in mice lacking circadian-related genes.^28,29^ However, the presence of an abnormal circadian rhythm has not been conclusively demonstrated in diabetic conditions.^30^ This is largely attributed to the absence of appropriate biomarkers for circadian rhythm.^31^ By utilizing D-alanine and its associated physiological process, gluconeogenesis, as biomarkers, we were able to visualize the abnormal circadian rhythm. The urinary level of D-alanine and gluconeogenic activity remained consistently elevated, while the blood level of D-alanine was persistently low. Both of these phenomena showed a loss of circadian oscillations. Notably, the increased urinary excretion of D-alanine likely contributed to the suppression of its blood level, as seen in awake mice. This finding was further corroborated in mice subjected to sleep disturbances.^13^ In cases of reduced eGFR, urinary excretion of D-amino acids, including D-alanine, is reduced.^20^ Consequently, the decreased blood level of D-alanine may be obscured because of the reduced GFR. Nevertheless, even under these conditions, urinary D-alanine and gluconeogenesis in the kidney serves as indicators of the abnormal circadian rhythm associated with diabetes.

D-Alanine possesses the ability to enhance gluconeogenesis even in the context of diabetes. In healthy kidney, D-alanine stimulates the expression of circadian clock transcriptional genes, which in turn promote the expression of gluconeogenic genes. As D-alanine exhibits a distinct circadian rhythm in the body, the activity of gluconeogenesis also follows a clear circadian rhythm. Additionally, D-alanine also acts as a substrate for pyruvate through its oxidation by D-amino acid oxidase;^13^ however, this effect is somewhat constrained because of the low concentrations of d-alanine present in the blood. In diabetic conditions, characterized by persistently elevated gluconeogenic activity, D-alanine continues to still has the potential to further enhance gluconeogenesis.

Under diabetic conditions, the activated gluconeogenesis contributes to elevated blood glucose levels. Therefore, D-alanine has the potential to increase blood glucose level upon administration of D-alanine. Conversely, D-alanine may also promote the normalization of blood glucose levels by correcting circadian rhythms. Between these two seemingly contradictory effects, D-alanine has demonstrated the ability to normalize glucose tolerance. Thus, the correction of circadian rhythms is crucial for regulating glucose metabolism through d-alanine. In addition to its role in glucose metabolism, D-alanine’s effect of circadian rhythm is important for the regulation of immunological processes. This is a potential connection between D-alanine and mitigation of viral infection exacerbation. Since the regulation of viral infections is particularly critical for patients with diabetes, D-alanine plays multiple roles foin the management of diabetes.

This study has several limitations. The primary parts of this study are based on rodent studies, necessitating further investigation into the effects in human subjects. The specific mechanisms of D-alanine under diabetic conditions were not explored, as the limitations of current scientific knowledge and technological capabilities. Nevertheless, the clear relationship between D-alanine and circadian rhythm demonstrated in this study is likely to pave the way to a new field of science for the clinical applications.

In conclusion, D-alanine is closely associated with diabetic conditions. The disruption of the circadian rhythm of D-alanine serves as a hallmark of diabetes, characterized by excessive urinary excretion of D-alanine. Although D-alanine retains the ability to upregulate gluconeogenesis in diabetic states, correction of circadian rhythm by d-alanine offers a potential in managing diabetes. The findings of this study hold significant implications for the future clinical application of D-amino acids.

## Data Availability

All data produced in the present work are contained in the manuscript

## Acknowledgments

We thank Hiroshi Imoto, Eiichi Negishi, Shoto Ishigo (KAGAMI Inc), and Yoko Tanaka for technical support. This study was supported by Japan Society for the Promotion of Science (22K194140) and Manpei Suzuki Diabetes Foundation.

